# On The Dynamics of Interstate Diffusion of Firearm Violence and Impact of Firearm Regulations

**DOI:** 10.1101/2021.11.15.21266358

**Authors:** Swagatam Sen, Anindya Sen, Ye Liu, Bisakha Sen

## Abstract

**Objectives:** Our purpose was to test the impact of firearm regulations on the firearm violence flow across US state borders. Further we assessed the spatial variations in these impacts across different regions with the goal of identifying state-groups that are especially vulnerable to cross-border firearm violence.

**Methods:** Incidence of firearm violence (2000-2017) has been modelled as an inhomogeneous diffusion process whose parameters depend on state firearm regulations. Firearm regulations measurement for a state accounted for all 14 law categories across 54 states since 1991 as per State Firearm Law Database. The effects of regulations and other covariates were estimated across all states.

**Results:** Six clusters of states were identified based on the variations of effects within and across those clusters. For 3 of these clusters the diffusive flow parameters were statistically significant. In all of these clusters the deterring effect of regulations on incidence and flow of crime was statistically significant.

**Conclusion:** The clusters can be assigned to 5 descriptive categories based on their roles in the flow of firearm violence – Source states, Transitive states, Destination states, Isolated issue states and Stable. It was found that flow of firearm violence indeed does follow a diffusive process for most categories of states. It has also been recommended that while in-state regulations are important to curb firearm violence flowing into Destination states, they are not adequate unless regulatory stringency is also applied to neighboring Source and Transitive States.

## Introduction

Injury and mortality related to firearm violence continues to be a major health concern for US. Firearm violence has been particularly severe over 2020-2021^1^, and response from the political leadership has followed a familiar pattern, with Democrat lawmakers calling for stronger gun regulations and Republican lawmakers opposing that idea. On June 23, 2021 President Biden’s administration announced their strategy to combat gun violence using funding from the Rescue Plan, which include measures to stem the spatial flow of firearms, including the illegal trafficking of firearms across state lines ^2^.

The associations between state-level firearm regulations on firearm violence has been well researched and documented^3-13^. Findings generally support negative associations between strength of firearm regulations and firearm violence, though effectiveness varies across sub-populations^7,14-17^. More recently, researchers have begun exploring links between state-level firearm regulations and firearm violence in neighboring states. Firearms are inherently highly transportable, thus allowing for relatively easy firearm migration and firearm trafficking across state borders^18^. Hence, a discrepancy between firearm-related policies in nearby states can lead to firearms being transferred from low-regulation states to high-regulation states, and potentially contribute to firearm violence in the high-regulation states^19,20^. For example, firearm shows in Nevada have been linked to short-term increases in firearm injuries in neighboring California^21^. Research using county-level and state-level data have shown evidence that firearms recovered in criminal investigations can frequently be traced to other states with less firearm regulation^22-24^, and that less regulation in nearby states are associated with higher firearm deaths in the home state. Further, not accounting for laws in neighboring states may spuriously reduce the strength of correlation between a state’s own laws and reduction in its own firearm deaths^13,25^.

The typical empirical approach in this literature uses time-series cross-sectional models, which employ area-specific and time-specific ‘fixed effects’ to reduce the influence of confounders such as area-specific socioeconomic and cultural characteristics, which are linked to both to firearm regulations and firearm violence. Such models essentially assume a ‘steady state’ ^26^, which does not account for how firearm violence evolves over time and spreads across states. Further, such models tend to obtain an “average effect” of firearm policies on forearm violence that is not confounded by state-specific factors. What this approach obscures is that states may not necessarily share a common pattern of how firearm violence evolves and moves in and around them.

In contrast, there exists a robust parallel literature on firearm violence that recognizes the dynamic and contagious nature of such violence, and model it as a diffusion process that follows distinct spatial and temporal patterns as it spreads through at-risk populations^27-31^. This has important policy implications. Static clustering of firearm violence emphasizes the need for place-based interventions that address underlying socio-economic-cultural challenges^32^, while diffusion models support interventions that aim to “interrupt” the transmission of violence at various points in the network^33^. This parallel literature with diffusion models have focused on cities and neighborhoods. However, it is logical to assume that similar diffusion processes may exist at the state level. Criminal activity, retributive violence, as well as social networks and illegal markets through which guns are transmitted across state-lines can evolve in response to changes in state regulations. Thus, there is a strong rationale for assuming that there exists a dynamic process wherein both the regulations and crime incidence patterns in a state can evolve alongside and based on those of its neighboring states^34^.

In light of the potential dynamic evolution of firearm violence propensity as well as relevant policy adoptions across states, and given evidence that gun-violence within smaller geographical areas like cities exhibit evidence of diffusion, it is important to challenge the steady-state assumption in the gun regulations literature. Our study helps fill this important gap in the literature.

## Methods

### Study sample

The study period is 2000-2017. All states of the US are included except Alaska and Hawaii because they are noncontiguous with other U.S. states. The District of Columbia is excluded because it has no applicable state laws in the State Firearm Law Database. The final analysis included 48 states.

### Outcomes

The outcome variable of interest is firearm death from all intent. Data on firearm deaths by state from 2000 to 2017 were extracted from the public-access Web-based Injury Statistics Query and Reporting System (WISQARS™) of the Center of Disease Control and Prevention (CDC)^35^

### State firearm policy

Information on state firearm law was obtained from the State Firearm Law Database^36^ developed by Siegel et al.^37^ This database tracks the presence of 134 law provisions in 14 categories across all 50 states for the period 1991 to the present. Examples of the categories are buyer regulations, dealer regulations, background checks, prohibitors for firearm purchase and possession, domestic violence-related firearm laws, “stand your ground” laws, concealed carry permitting laws, assault weapons regulations, firearm trafficking laws, and restrictions on places where firearms may be carried. Each of the 134 provisions was coded as being either present (1) or absent (0) for each state during each year. Laws were coded based on their year of implementation. The total number of laws by state and year ranged from 0 to 134.

### Other Covariates

Based on existing literature, we considered several additional covariates which are listed below^6,7,13,23^. Population size, proportion of age 65 or older, race, unemployment rate, poverty-rate, and the proportion age 25 or older without a high-school diploma were obtained from the US Census Bureau for 2000-2017 ^38^. Property crime rates were obtained from the Federal Bureau of Investigation’s summary reporting system via Crime Data Explore^39^ as a proxy for the propensity for crimes in the state. The per capita number of licensed firearm-dealers, obtained from U.S. Bureau of Alcohol, Tobacco and Firearms^40^, and the percentage of the hunting-license holder of the state’s population, obtained from the National Survey of Fishing, Hunting and Wildlife-Associated Recreation^41^, were included as proxy controls for household firearm ownership, following prior studies^7,42,43^. As a further proxy-measure for state sentiment towards firearm-control, the vote share differences between the Republican and Democratic presidential candidates in each presidential election year within our study period^44^ were included, and extrapolated for years between presidential elections.

### Empirical Method

We use a type of simple diffusion process^45^ to model the changes in firearm crimes and how changes in regulation in and around a state impacts that process, using an approach similar to Cohen et al.^28^ and Simmons et al. ^46^ to study diffusion of policies around human trafficking.

The key assumption in a pure diffusion process is that rate of flow through a given surface element of unit area is proportional to relative difference between the densities on two sides of that surface. This is formalized by the Fick’s law ^47,48^

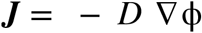

Wherein, in the present context, ϕ represents incidence of firearm death in a given state in a year, and *J* is the rate outward flow of firearm death. In this article, we assume the diffusion process to take place in a non-homogeneous medium where coefficient *D* = *D*(***x)*** varies across states.

Using Diffusion models to understand the flow of firearm violence across state borders also allows to investigate the differential nature of such flows across different regions and their mutual interactions within the broader network.

The inhomogeneous stochastic evolution model for the incidence of gun-related violence yields a discretized version that is in the form of a linear model.

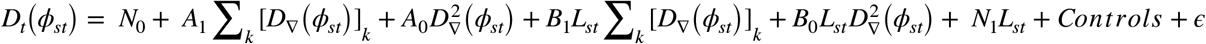

The variables of interest in the model are listed below.

*ϕ*_*st*_: Incidence rate of gun-related crimes in state s and year t

*D*_*t*_(*ϕ*_*st*_): *Target -* Calculated as 1-step increment in rate of firearm-related violence over time with years counted as units of time.

*D*_∇_(*ϕ*_*st*_): *Diffusion-Index-1 -* Difference between firearm violence rate a state versus proximal states.

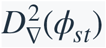: *Diffusion-Index-2 –* The difference in difference between firearms violence in a state versus proximal states.

*L*_*st*_: *Regulations –* This variable quantifies the strictness of firearm control regulations within a state.

*Diffusion-Interaction-1 and Diffusion-Interaction-2* - The regression model also incorporates interaction terms between *the Diffusion* Indices *and Regulations*, represented as products of the variables.

*Controls -* To account for residual variability present in the crime rates related to firearms, which are extrinsic to the effects of regulations, we included a number of control factors into the model - 1) Population density in the state, 2) Poverty Index, 3) Unemployment rate, 4) Percentage of population over 65 years age, 5) Percentage of White population, 6) Percentage of Hunting license holders (as proxy for gun ownership levels) and 7) Property crime rates (as proxy for general crime propensity in the state) out of which only 1, 2, 4 and 5 turned out to be significant factors for one or more states or state groups during the study period.

The details of the calculation procedures for the variables of interest are presented in Appendix B.

The effects of the factors together define the dynamics of the process and help to answer key questions around it. If there is an impact of gun regulations on new gun sales rate, then we should find effects of *Regulations* to be significantly non-zero. Similarly a significant effect of *Diffusion-index-1* or *Diffusion-index-2* for a state or cluster of states would indicate substantial diffusion of crime - either incoming or outgoing - happening across the borders of such state(s). An impact of gun regulations on such a flow of crimes would manifest in non-zero values of either *Diffusion-Interaction-1* or *Diffusion-Interaction-2* or both.

## Results

The study identified 6 distinct groups or clusters of states based on how firearm deaths flow across the borders of these states (*see Table 1)*. These states are not necessarily contiguous or similar in their crime incidence rate and regulations, even though it is generally observed that state groups with more regulations tend to have fewer incidences of firearm related deaths (*See*

*Figure 1)*. Cluster 1 states have applied highest number of regulations (average regulations = 62) with the effect that they enjoy significantly lower firearm death rate (incidence rate = 0.006%). However, such patterns cannot be generalized. Cluster 2 states have historically adopted much stronger regulatory regime (average regulations = 28) compared to that of Cluster 4 states (average regulations = 16) even though both the groups observe same propensity for firearm deaths (incidence rate = 0.011%).

*Table 2* reports the results of the final models for each of the groups. This includes estimated effect and statistical significance of Regulations, Diffusion Indices and effects of regulations on diffusion indices, as well as control covariates. Within each state clusters *Regulations* have a consistently negative association with firearm deaths. In four out of six groups (0,1,2 and 3), this effect is significantly prominent (for example, Cluster 1: weight of *Regulations* = - 0.59434, p-value=3.64E-08), while in other states the deterrence effect, while observed, is not significant.

In three out of the six groups – groups 0, 2 and 3 – traces of diffusion are evident from the significance of the estimates for *Diffusion-Index-1* and *Diffusion-Index-2*. In fact these two estimates are of opposite sign for all the clusters (*See Table 2*) reconfirming the diffusive nature of the underlying process. Henceforth we would define groups (and states within them) as *Inwardly Diffusive* if the weight estimate for *Diffusion-Index-2* is negative, and *Outwardly Diffusive* if the said estimate is positive.

Estimates of *Diffusion Indices* exhibit differences in crime migration patterns across 6 state groups.

1. Cluster 1 and 5 states have statistically insignificant diffusion index (p-value = 0.67 and 0.44 respectively) which indicates that there are no significant diffusion of crime across the borders of these states rendering them diffusively neutral.

2. The states in Cluster 0 and 2 demonstrate strong *Inward Diffusive* pattern of firearm death flow (**Cluster 0 :** weight of *Diffusion-Index-2* = −2.91972, p-value = 2.09E-16; **Cluster 2 :** weight of *Diffusion-Index-2* = −1.79533, p-value = 2.35E-11)

3. Cluster 3 states are strongly *Outward Diffusive* (weight of *Diffusion-Index-2* = 1.654576, p-value = 0.000757) while Cluster 4 states show mild traces of outward diffusion (weight of *Diffusion-Index-2* = 1.654576, p-value = 0.019628)

The estimates of *Diffusion-Interaction-1* and *Diffusion-Interaction-2*, in general indicate a deterring effect of regulations on the extent of diffusive flow of firearm deaths in state groups where significant traces of diffusion were observed. We can quantify the deterrent impact of regulations on the diffusion flow through a state, by normalizing the effect of *Diffusion-Interaction-2* by that of *Diffusion-Index-2*,

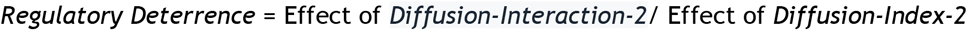

*Regulatory Deterrence* while observed for all clusters where Diffusion is present, is only significant in Clusters 0 and 2 (*See* ***Table 2;*** *Cluster 0 =-1*.*27, Cluster 2 = −0*.*87)* which are the same clusters that we observed to be highly vulnerable to inward diffusion of firearm death from across neighboring states.

## Discussion & Conclusion

The present work has targeted to understand the flow of dynamics of firearm related crimes across the states of USA (excluding Hawaii and Alaska due to lack of contiguity). That flow has been modelled as an inhomogeneous diffusion process.

Results showed that the states can be grouped into 6 broad clusters with significantly different firearm death incidence, regulations, nature and extent of firearm death diffusion, and impacts of regulations on it. Based on the type of diffusion, these clusters can further be classified into 5 descriptive categories that we name as follows (*See Figure 2)*.

- ‘Source’ States (Cluster 3) - These are mostly Northern states in the Cluster 3 and are characterized by high within-state crime rates as well as very significant outward diffusion of crime. It has been observed that firearm deaths in these states could be controlled by increased regulations, but the states have the lowest number of firearm regulations. There is a considerable policy opportunity presented by these states to tighten the regulations which would assist in controlling firearm violence in these states as well as the diffusion across borders.
- ‘Transitive’ States (Cluster 4) - States in Cluster 4 which are mostly located in a geographic belt between Source states and the Destination states. These transitive states benefit from a relatively low crime rate despite fewer regulations. However, these states show mild outward diffusion - a characteristic resulting from net effect of inward and outward migration of crime through these states and seemingly unaffected by increased regulations. These insights point to the conclusion that these states, even though not the source of flow, acts as propagator of crimes to the destination states.
- ‘Destination’ States (Clusters 0 and 2) - Most of these states from Cluster 0 and 2, are located along and towards the east coast. These suffer from moderate-to-high firearm related crime rates. Some of the states in Cluster 2 have very high number of regulations within state. However, the impact on crime rate has not been commensurate, as these states continue to be the victim of mostly cross-border firearms and crimes migrating from the Source and Transitive states.
- ‘Stable’ States (Cluster 1) - Towards the north-east coast of US and around New York, there are a cluster of states that demonstrate the best scenario when it comes to managing firearm violence. Despite the high population density, these states have historically had put in place a strong regulatory framework which has resulted in the lowest firearm related crime rate by far. Further, these states appear to be largely immune to cross-border migration of crime as they are mostly surrounded by ‘Destination’ States with strong regulations.
- Isolated Issue States (Cluster 5) - The states in Cluster 5 either share an international border or are too close to it. These states exhibit a high rate of firearm violence which is in part triggered by some of the lowest number of regulations within states. However, these states are diffusively neutral, indicating that these states represent concentrated hot-spots of crime influenced by the international border rather than the broader flow of crime impacting the other states.

In summary, we conclude that gun-related crime does not only have diffusive effect on neighboring states or that more lenient regulations in a neighboring state can have negative influence on gun violence deterrence, but such flow of crime has a distinct route which has been stable over the years.

It is quite evident from the analyses that while in-state regulations do matter to some extent to control firearm violence, for many of the states one of the primary driver for crime is the inward migration across borders with neighboring states, particularly from some of the identified source states. Clearly primary policy recommendation should be to strengthen firearm regulations in the Source states.

There is also a need for broader regulations in Isolated Issue states, particularly because of their proximity to international borders. Since the violence pattern seems to be localized, a timely intervention can help to stop the spread in future.

### Limitations

This study has limitations. We have not considered specific types of firearm regulations adopted by various states, which may potentially further explain why some of the states have had more success through increased regulations as opposed to others. Based on the analyses, future studies should analyze and compare regulations used by the Stable states to a great effect, as best practices for other states. Further, we have not segregated violence between homicides and suicides and have reserved that for a more detailed subsequent study. In addition, this study design could not control for state specific unobserved factors (for example, attitude/cultural modality/political orientation) that may potentially confound the relationship between firearm policies and firearm deaths.

In conclusion, within the stated limitations of the work, we have been able to add to the literature by providing further insights into the dynamics of firearm related violence and how it migrates across state borders. We believe this work helps identify some of the concerning corridors of migration and have demonstrated how such flows can impact overall evolution of firearm related crimes across all states.

## Data Availability

All data produced in the present study are available upon reasonable request to the authors

## Appendix A

### A Stochastic Model for Gun Trafficking

The most generic form of diffusion process would be

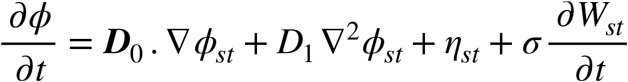

where *η* is the rate of gun sales, *W* is a Wiener process and *D*_1_, ***D***_0_ are diffusion coefficient and diffusion gradients. Furthermore while *s* denotes the state index, ∇ in the equation represents partial derivatives along actual physical separation across states.

For our present purposes, we’d specifically investigate -

1. Is there an impact of Gun regulations on new gun sales?

2. Is the gun migration across states influenced by respective states’ gun regulations?

In other words, we can explore the functional relationships of *D*_1_, ***D***_0_ and *η* with the number of gun regulations (*L*_*st*_). Furthermore in the present work we have only explored simple linear relationships between these quantities, with a clear scope to explore other types elsewhere.

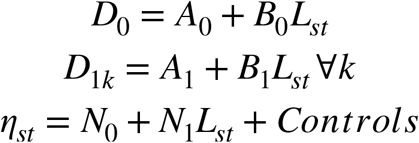

It is to be noted here that we are implicitly assuming the diffusion gradients to be symmetric i.e. *D*_1*k*_ are the same ∀*k*. It’s a simplifying assumption at this stage and can be reasonably challenged in subsequent work.

With these assumptions, we can arrive at the final for the model,

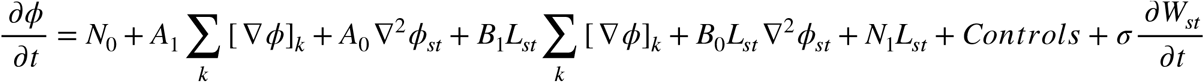

In this form, the model reduces to linear regression model for 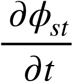 with the predictors on the right hand side if one can calculate the relevant quantities.

## Appendix B

### Description of model effects and control covariates

*ϕ*_*st*_: Incidence rate of gun-related crimes in state s and year t

*Dt*(*ϕst*): *Time-increment-in-Firearm-Crimes -* Calculated as 1-step increment in rate of firearm-related violence over time with years counted as units of time.

*D*_∇_(*ϕ*_*st*_): *tate-Difference-in-Firearm-Crimes-1 -* Difference between firearm violence rate a state versus proximal states. Proximal states have been identified based on whether the distance from the concerned state is less than 500 miles.

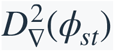: *State-Difference-in-Firearm-Crimes-2 –* The difference in difference between firearms violence in a state versus proximal states. Mathematically, this corresponds to the discretized version of the second order spatial derivative of *ϕ*_*st*_ in the diffusion model.

*L*_*st*_: *Regulations –* This variable quantifies the strictness of firearm control regulations within a state in the form of total number of firearm regulations.

*Interaction Terms* - The regression model also incorporates interaction terms between the variables listed above, represented as products of the variables.

*Controls -* To account for residual variability present in the crime rates related to firearms, which are extrinsic to the effects of regulations, we included a number of control factors into the model - 1) Population density in the state, 2) Poverty Index, 3) Unemployment rate, 4) Percentage of population over 65 years age, 5) Percentage of White population, 6) Percentage of Hunting license holders (as proxy for gun ownership levels) and 7) Property crime rates (as proxy for general crime propensity in the state).

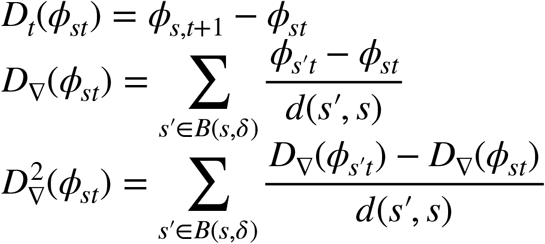

In these calculations, we have used a state-to-state distance function *d* which is calculated based on centroids of each state. Using this distance we have also defined a ‘neighborhood’ for each state *s* as *B*(*s, δ*) = {*s*′| *d*(*s*′, *s*) < *δ*}, where *δ* is an arbitrary proximity parameter.

